# Social deprivation and profile of referrals and disability in paediatric chronic pain: a cross-sectional study

**DOI:** 10.1101/2020.02.24.20027037

**Authors:** Eveline Matthews, Maeve Muldoon, Norma O’Keeffe, Kevin McCarthy

## Abstract

Social deprivation is associated with a higher prevalence of chronic pain in children and an under-representation in specialist paediatric chronic pain programs. This study is a retrospective, cross-sectional analysis of referrals to the National Paediatric Complex Pain Service in Ireland to assess if there is any link between social deprivation and pattern of referrals. Secondary objectives included determining any additional differences between deprivation categories and pain characteristics, parental pain catastrophizing, and pain-related disability, including sleep quality and school attendance. We recorded data on 289 children referred to the NPCPS between February 2016 and November 2019. Social deprivation was assessed using the Pobal HP Deprivation Index, which is based on Irish national census data. The results of the primary analysis showed referrals have a normal distribution across the deprivation index, although the disadvantaged group had a longer duration of pain at time of first clinic review (p=0.01). Secondary analyses showed deprivation is associated with higher levels of parental pain catastrophizing (p=0.0016), most significantly for helplessness (p=0.0009), and higher use of screens at bedtime (p=0.002) with longer sleep onset latency (p=0.04). However, there were similar rates of sleep disturbance, school attendance, social interference across the deprivation groups. These findings may indicate inequities in access or differences in service utilization for children from socially disadvantaged families. The differences in sleep hygiene and parental pain catastrophizing may indicate the need to screen for these potential mediators of treatment outcome, which may require targeted intervention when present, in order to harmonise treatment responses across deprivation grades.

**Research in context:** *Evidence before this study:* Social deprivation is associated with an increased risk of chronic pain in childhood, however children from deprived areas may be under-represented in specialist paediatric pain centres. This is be due to inequities in access to services and also differences in healthcare utilisation, possibly due the cognitive burden of poverty.

*Added value of this study:* This study found that while referrals were normally distributed across social deprivation grades, children from the disadvantaged group had pain for longer prior to their first clinic review. This may indicate differences in healthcare utilisation by disadvantaged families. Additionally, we found differences in sleep hygiene and parental pain catastrophizing, which are both of these are known mediators of pain chronicity.

*Implications of all available evidence:* Due to potential differences in healthcare utilisation across deprivation grades, clinical pathways might need adaptation for disadvantaged families for greater efficacy. Targeted interventions that address sleep hygiene and parental catastrophizing might also be considered early in a treatment pathway for disadvantaged families, perhaps with more scheduled follow-up, to harmonise treatment responses across deprivation grades.

## Introduction

Socioeconomic deprivation is a determinant of mortality from chronic illness^i^. This effect is mediated through the magnification of known unhealthy lifestyle factors, such as physical inactivity, in deprived populations^ii^. In addition to all-cause mortality, multi-morbidity occurs at a younger age in those living in areas of high social deprivation and most frequently involves a combination of physical and mental health problems, with depression and pain remaining among the most common conditions across all age groups^iii^. Chronic pain is not only more common in socially deprived areas, it is also associated with greater pain intensity and pain–related disability, and the economic burden of pain further compounds the initiating and maintaining factors of deprivation^iv^. The precise mechanisms mediating the effect of deprivation on chronic pain in adulthood are likely complex and multifactorial^v^.

Evidence exists of longitudinal relationships between social deprivation and pain in childhood and chronic pain in adulthood. The longitudinal 1946 British birth cohort study found that abdominal pain, poorest care in childhood, and poorer maternal health were associated with an increased risk of persistent low back pain in both sexes in early- and mid-adulthood^vi^. Links between social deprivation and persistent pain in childhood are also emerging^vii,viii^. Household income and socioeconomic status is also a determinant of healthcare utilisation^ix^, referral to paediatric pain clinics^x^ and to intensive paediatric pain treatment^xi^.

The primary objective of this study was to examine any influence of social deprivation on referrals to a national paediatric chronic pain service. Additional outcomes of secondary interest included any differences in other variables such as pain intensity and duration, sleep, school attendance, and parental pain catastrophizing, all of which are recorded at first clinic visit.

## Methods

### Participants

Following institutional ethical approval, we performed a retrospective, cross-sectional analysis of referrals to the National Paediatric Complex Pain Service (NPCPC) in Ireland. Referrals to this service are children and young people in the Republic of Ireland with chronic pain. It includes data from 289 children and young people referred to the service between February 2016 and November 2019.

### Complex Pain Assessment

Upon initial referral to the pain service, children and their parents complete a complex pain assessment form, which records pain intensity and duration, and parental estimates of children’s pain intensity and coping ability, each of which were recorded on an 11-point numerical rating scale (NRS). Factors related to sleep quality are also recorded, such as use of digital devices immediately before bedtime and self-reported sleep onset latency (<30 minutes, 30-60 minutes, >60 minutes) and pain-related sleep disruptions, asked as percentage of wakings that are due to pain. The impact of pain upon school attendance is recorded by self-reported number of days missed and percentage school attendance. Pain interference with social activities is a recorded as a self-reported 11 point 0-10 NRS “0- no interference” to “10 - completely interferes”. We also administer the Pain Catastrophizing Scale-Parent (PCS-P)^xii^, as in the context of paediatric pain, high levels of parental pain catastrophizing has been linked to poor outcomes for children with pain, likely through an influence on child pain catastrophizing^xiii^.

### Deprivation Index

Social deprivation was assessed using the Pobal HP Deprivation Index. This index provides a method of analysing comparative affluence and deprivation by geographical area derived from data from the 2016 National Census of Ireland. The Deprivation Index is based **on** geographic units called small areas (SAs), each of which are uniform in size, with a mean of approximately 100 households, and are relatively homogenous in social composition. The Pobal HP Deprivation Index is based on three dimensions: demographic profile, social class composition, and labour market situation. The census data used as indicators of each of these dimensions includes percentage population change, mean number of persons per room, level of education, percentage of households headed by unskilled workers, and unemployment rate. The Relative Index Scores are specific to that census wave and are rescaled to have a mean of zero and a standard deviation (SD) of ten. The scores approximately follow a normal distribution in eight categories from ‘extremely affluent’ (>3SD above the mean), to ‘extremely disadvantaged’ (>3SD below the mean). The Relative Index Score and corresponding deprivation category for each participant were determined by entering their postal zip code into the Pobal online portal. For the between-group analyses, we used cut-offs of >1SD above the mean as ‘affluent’, <1SD above or below the mean as ‘average’ and >1SD below the mean as ‘disadvantaged’.

### Hypothesis

The primary objective of this analysis was determining whether social deprivation influenced access to care. Based on our clinical experience and other previously published work^x^, our hypothesis was that we were seeing more patients from affluent backgrounds than from disadvantaged backgrounds. To do this, we tested whether the total referrals followed a normal distribution across deprivation categories. We also looked for between-group differences in the duration of pain and first attendance at clinic.

Secondary objectives included determining any additional between-group differences in factors associated with pain, such as age and sex, pain intensity, sleep hygiene, pain impact upon function, such as sleep quality, school attendance and social activities, and parental estimates of pain intensity, child pain coping and parental pain catastrophizing.

### Statistical Analysis

Datasets were tested for a normal distribution using the Anderson-Darling test and nonparametric tests were used where appropriate. For between-group differences, we used a one-way analysis of variance (ANOVA) or Kruskal-Wallis test for continuous variables and a chi-squared test for categorical variables. Data is presented as median and interquartile range (IQR) unless otherwise stated. Significance was set at alpha =0.05 with correction for multiple comparisons on post hoc testing.

## Results

### Participants

We retrieved assessment sheets on 289 patients referred to the paediatric chronic pain service between February 2016 and November 2019. Patients (34% male) had a median age of 13 years (IQR 11-15 years) and had median average pain scores of 7 on a 0-10 NRS (IQR 5-10). The median duration of pain was 24 months (IQR 8-49.5 months).

### Access to Care

An Anderson-Darling test indicated that the 289 referrals over four years to the NPCPS [Figure 1] followed a normal distribution across deprivation categories, A_2_=0.535, p=0.11. There was however a significant difference in the median time from onset of pain to time of first review between deprivation groups, Kruskal-Wallis 8.7, p=0.013; specifically between middle (median 22 months, IQR 6.0-44.5 months) and disadvantaged (median 36 months, IQR 16.5-60.5 months) groups, Dunn’s multiple comparison test, mean rank difference −35.7, p=0.01.

**Figure 1:**
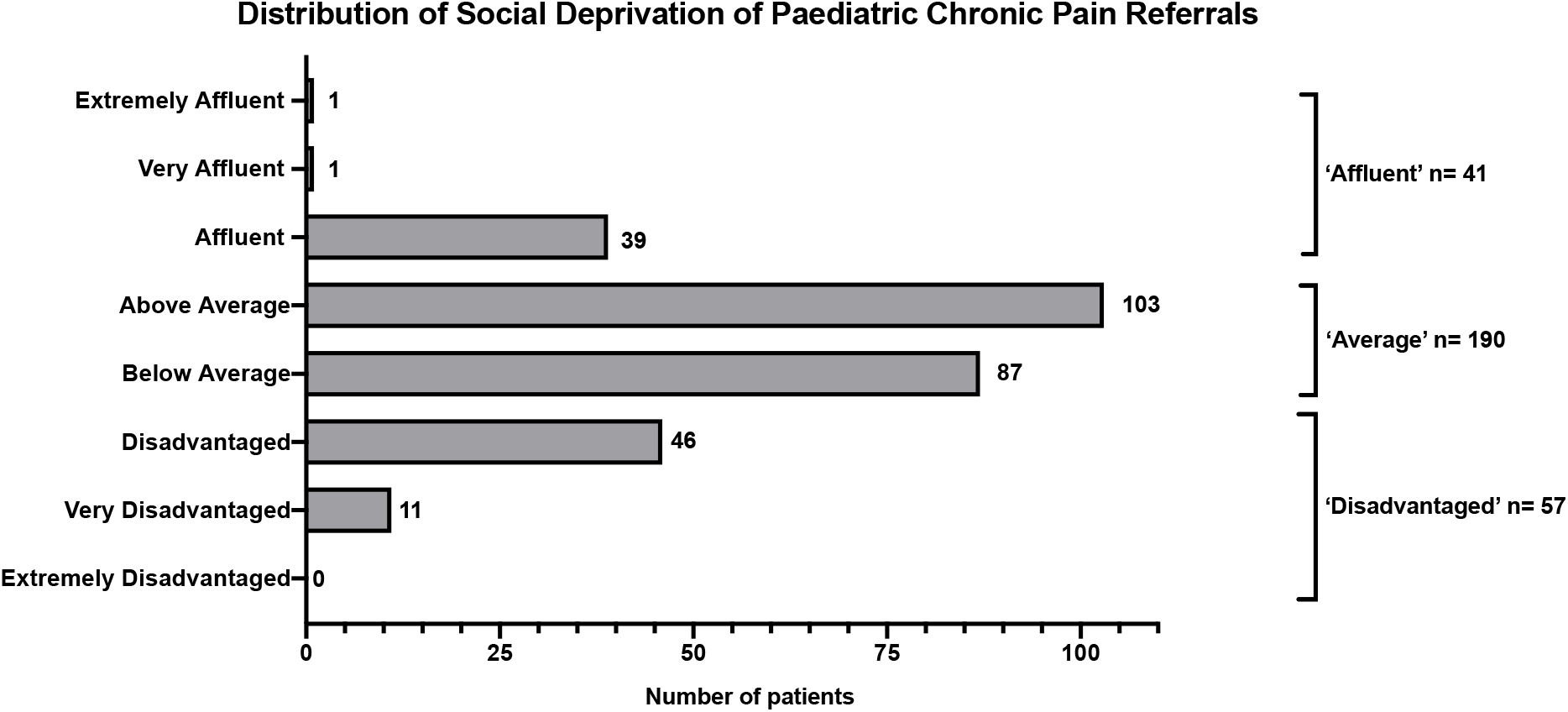
Distribution of Relative Index Scores, which are rescaled to a mean of zero and standard deviation (SD) of 10. A total of 289 records were available to analyse. For between-group analyses, the eight deprivation categories were regrouped into affluent (>1SD above the mean), average (<1SD above or below the mean) and disadvantaged (>1SD below the mean).

**Figure 2:**
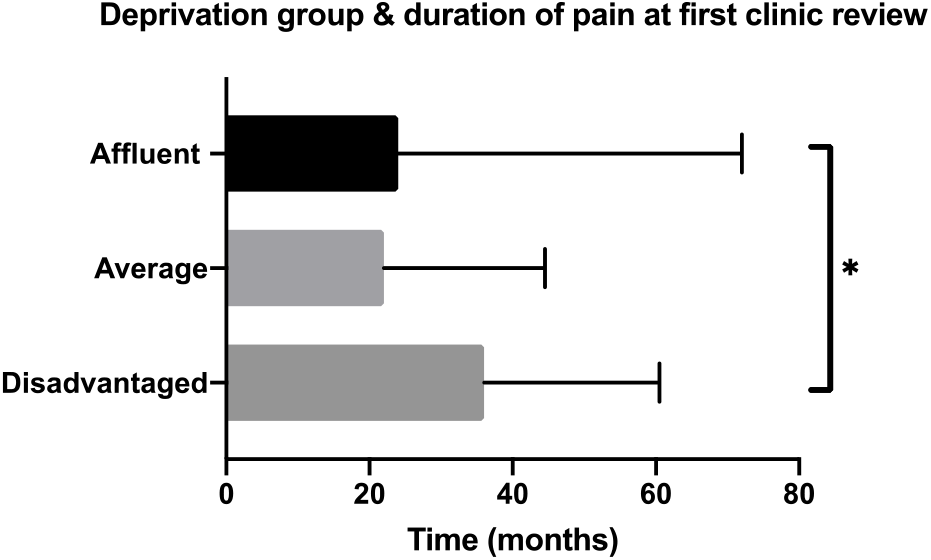
Disadvantaged group have pain for longer by time of first review. Median time from onset of pain to time of first review between deprivation groups, Kruskal-Wallis 8.7, p=0.013; specifically between middle (median 22 months, IQR 6.0-44.5 months) and disadvantaged (median 36 months, IQR 16.5-60.5 months) groups, Dunn’s multiple comparison test, mean rank difference −35.7, p=0.01.

**Figure 3:**
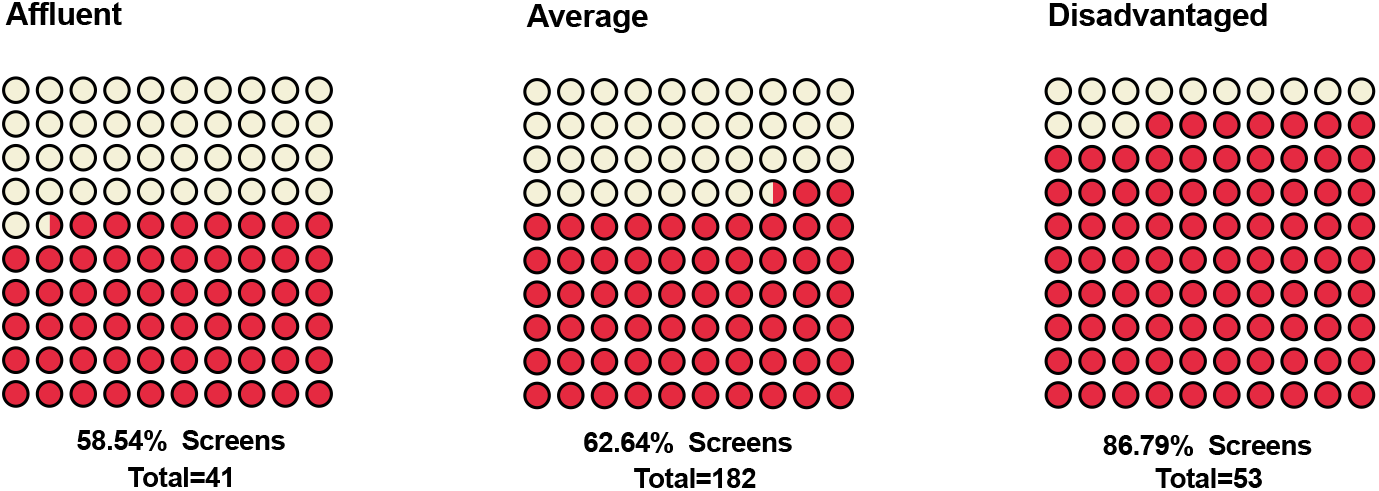
Deprivation group and use of screens before bed. Overall, 65% of children admitted to using screens or digital devices right before bed, which varied significantly between groups (58.5% of affluent vs. 87% of disadvantaged), Chi-squared *x2* (2,269)=12.21, p=0.002.

**Figure 4:**
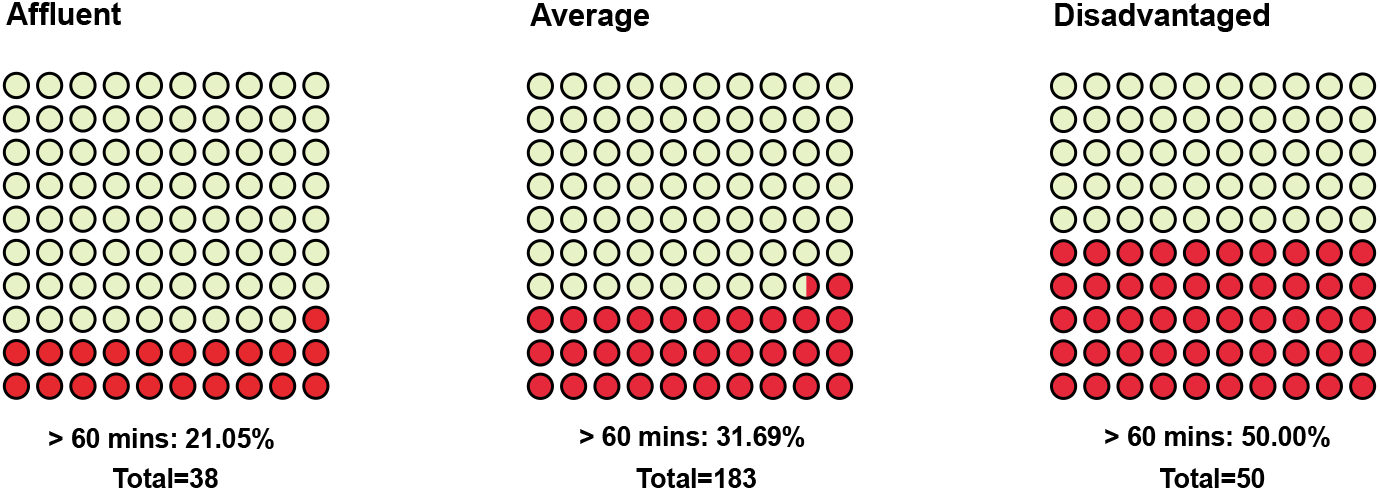
Deprivation group and prolonged sleep latency. The proportion of children reporting that it took over 60 minutes to fall asleep was significantly different between groups (21% of affluent vs. 50% of disadvantaged), Chi-squared *x2* (4,271)=10, p=0.04.

**Figure 5:**
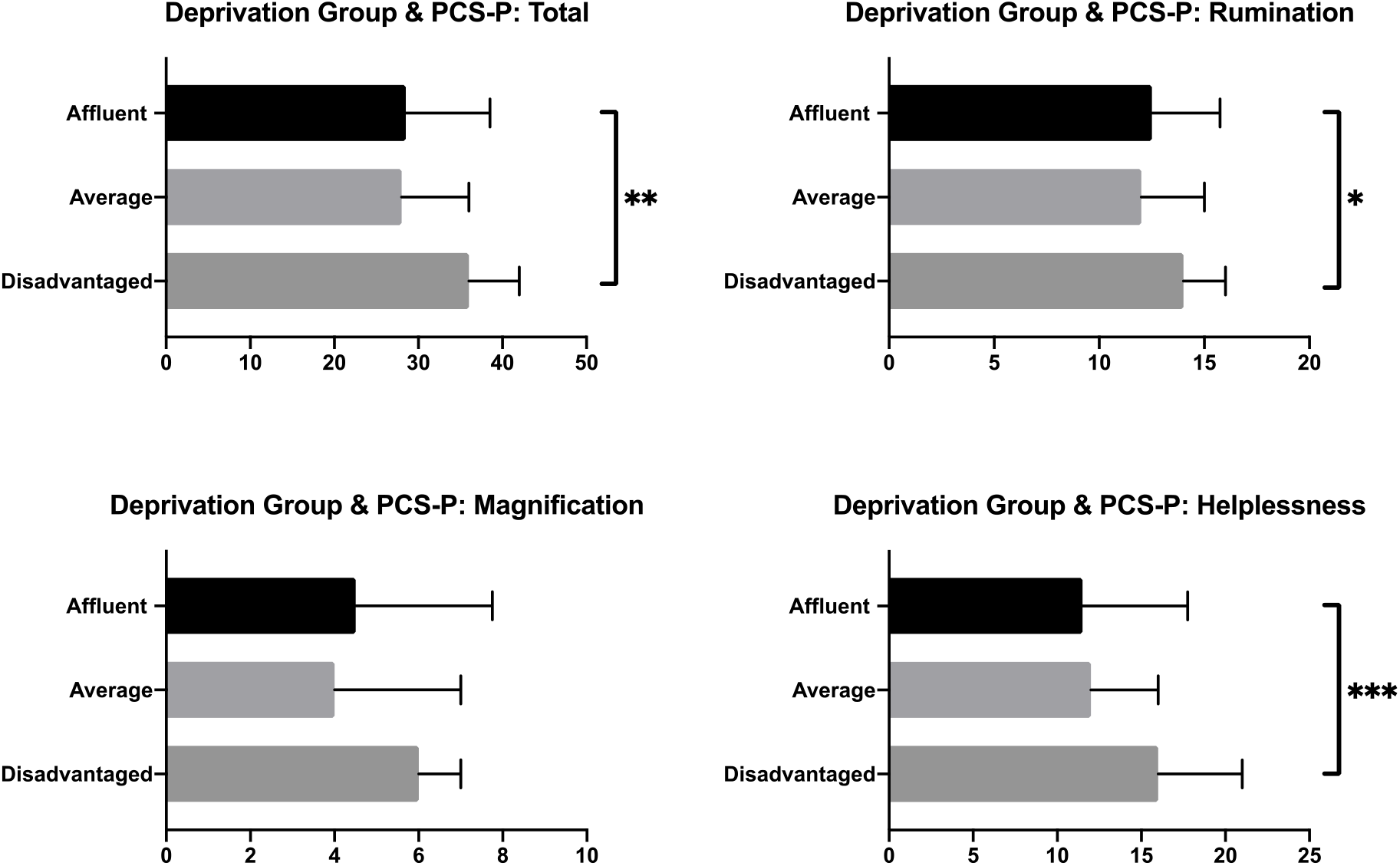
Deprivation groups and parental pain catastrophizing (PCS-P) scores. PCS-P median total scores were significantly different between groups, Kruskal-Wallis 12.81, p=0.0016. On Dunn’s multiple comparison test, the difference was between the disadvantaged group and both affluent (mean rank difference, −41.23, p=0.04) and average (mean rank difference −43.73, p=0.001) groups. On examining between-group differences of each subscale of the PCS-P, the rumination (Kruskal-Wallis 8.8, p=0.01) and helplessness (Kruskal-Wallis 13.9, p=0.0009) subscales were significantly different across groups and on post hoc testing of each difference it was the disadvantaged group that was different from one or both of the other two groups.

### Pain scores

Across the three deprivation groups, there was no difference in the medians of the lowest pain scores, Kruskal-Wallis 4.37, p=0.11, average pain scores, Kruskal-Wallis 0.084, p=0.95, or highest pain scores, Kruskal-Wallis 2.82, p=0.24.

### Age & sex

There was no difference in median age in years across the three groups, Kruskal-Wallis 5.43, p=0.06. The proportion of male patients was 34% of the total referrals and a Chi-squared test of independence showed no difference between groups, *χ*_*2*_(2, 288)=0.73, p=0.69.

|  |  | Affluent | Average | Disadvantaged | KW p-value |
| --- | --- | --- | --- | --- | --- |
| Pain Intensity 0-10 NRS | Average | 7 (6-8) | 7 (5-8) | 7 (5-8) | 0.95 |
|  | Highest | 9 (8.5 – 10) | 9 (8-10) | 9 (8-10) | 0.24 |
|  | Lowest | 3 (2-6) | 4 (3-6) | 5 (30) | 0.11 |
|  | Duration (mths) | 24 (9 – 72) | 22 (6-44) | 36 (16-60) | 0.01 |
| Age (yrs) |  | 14 (11-15) | 13 (11-15) | 14 (12-16) | 0.06 |
| Sex | Male (%) | 16/41 (39%) | 61/189 (32%) | 20/57 (35%) | 0.69 |

### Sleep hygiene

The median weekday bedtime (21.00 hours, IQR 21.00-22.00 hours) was not significantly different between groups, Kruskal-Wallis 2.89, p=0.23. The median weekend bedtime (22.00 hours, IQR 22.00-23.00 hours) was also not significantly different between groups, Kruskal-Wallis 4.71, p=0.09. Overall, 65% of children admitted to using screens or digital devices right before bed, which varied significantly between groups (58.5% of affluent vs. 87% of disadvantaged), Chi-squared *χ*_*2*_ (2,269)=12.21, p=0.002. With regard to self-reported sleep latency, the proportion of children reporting that it took over 60 minutes to fall asleep was significantly different between groups (21% of affluent vs. 50% of disadvantaged), Chi-squared *χ*_*2*_ (4,271)=10, p=0.04.

### Pain & function: sleep, school, social

The impact of pain upon sleep quality was assessed by asking whether the number of wakings from sleep that were due to pain was less than, equal, or more than 50% of all sleep disturbances. The frequency of pain-related sleep disturbance was not different across social deprivation groups, *χ*_*2*_ (4, 227)=6.3, p=0.177. School attendance was reported by parents as either number of days missed or percentage attendance and there were no significant differences between deprivation groups, Kruskal-Wallis 5.17, p=0.07 for days missed and Kruskal-Wallis 3.0, p=0.21 for percentage attendance. Pain interference with social activities did also not differ significantly between groups, Kruskal-Wallis 1.88, p=0.38.

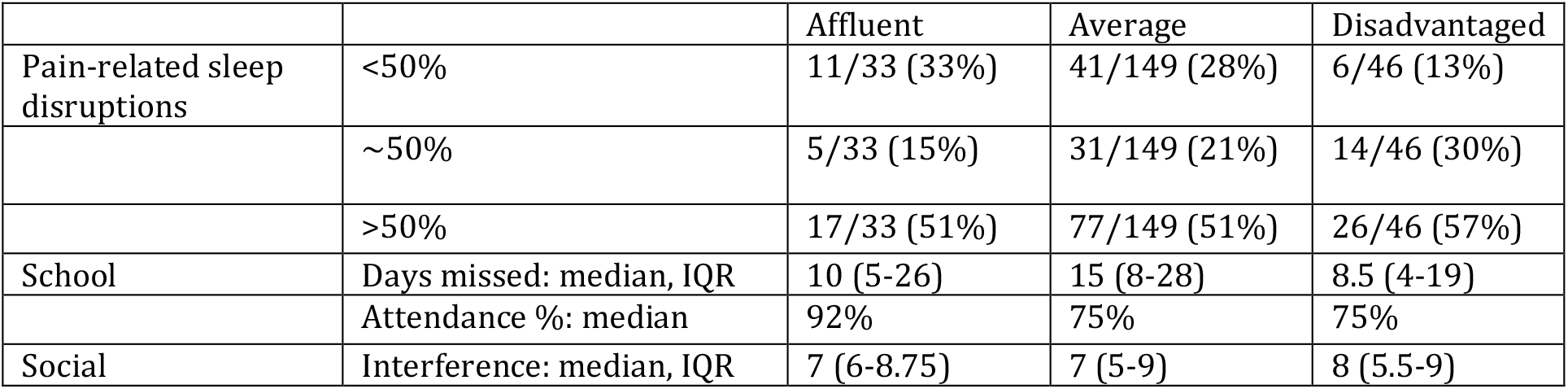

|  |  | Affluent | Average | Disadvantaged |
| --- | --- | --- | --- | --- |
| Pain-related sleep disruptions | <50% | 11/33 (33%) | 41/149 (28%) | 6/46 (13%) |
|  | ~50% | 5/33 (15%) | 31/149 (21%) | 14/46 (30%) |
|  | >50% | 17/33 (51%) | 77/149 (51%) | 26/46 (57%) |
| School | Days missed: median, IQR | 10 (5-26) | 15 (8-28) | 8.5 (4-19) |
|  | Attendance %: median | 92% | 75% | 75% |
| Social | Interference: median, IQR | 7 (6-8.75) | 7 (5-9) | 8 (5.5-9) |

### Parental estimates: pain intensity & pain coping

Parental estimates of average pain intensity, KW 0.85, p=0.65, and of their child’s ability to cope with pain, KW 1.46, p=0.48, were not significantly different between groups.

|  |  | Affluent | Average | Disadvantaged |
| --- | --- | --- | --- | --- |
| Parental estimates | Average pain 1-10 NRS (median, IQR) | 7 (6-8) | 6.5 (5-8) | 7 (5-8) |
|  | Coping 0-10 NRS (median, IQR) | 6 (4-8) | 6 (4-7) | 6 (5-8) |

### Parental pain catastrophizing

PCS-P median total scores were significantly different between groups, Kruskal-Wallis 12.81, p=0.0016; on Dunn’s multiple comparison test, the difference was between the disadvantaged group and both affluent (Mean rank difference, −41.23, p=0.04) and average (Mean rank difference −43.73, p=0.001) groups. On examining between-group differences of each subscale of the PCS-P, the rumination (Kruskal-Wallis 8.8, p=0.01) and helplessness (Kruskal-Wallis 13.9, p=0.0009) subscales were significantly different across groups and on post hoc testing of each difference it was the disadvantaged group that was different from one or both of the other two groups

|  |  | Affluent | Average | Disadvantaged | KW p-value |
| --- | --- | --- | --- | --- | --- |
| PCS-P | **Total** | 28.5 (19.25- 38.5) | 28 (20-36) | **36 (29-42)** | **0.0016** |
|  | **Rumination** | 12.5 (10-15.75) | 12 (10-15) | **14 (12-16)** | **0.0120** |
|  | Magnification | 4.5 (3-7.75) | 4 (2-7) | 6 (4-7) | 0.0728 |
|  | **Helplessness** | 11.5 (6-17.75) | 12 (7-16) | **16 (12-21)** | **0.0009** |

## Discussion

Our hypothesis was that the service was seeing greater numbers of children from affluent families and households. However, in our sample, referrals were normally distributed across deprivation grades. There were no differences between the deprivation groups for age, sex, pain scores, parental estimates of pan and pain coping, or for functional outcomes such as sleep disturbance, school attendance or interference with social activities. The disadvantaged group differed significantly for duration of pain prior to first review, use of screens before bedtime, prolonged sleep latency, and higher levels of parental pain catastrophizing.

Although different countries utilize different measures of socioeconomic status, a relationship between affluence and household income with access to specialized paediatric pain clinics has been previously described^xxi^. In addition, these studies also suggested that greater distance from the clinic was a determinant of referral and attendance. Ireland is a geographically small country which might mitigate the impact of travel times somewhat. Irish healthcare is a two-tier mix of private health insurance and public state-funded care, with approximately 45% of the population having some form of private insurance^xiv^. The NPCPS is a national service available to all, and Irish patients have the option opting in or out of using their insurance for different episodes of care. Therefore, we felt this would not be a meaningful variable to measure. However, insurance status might account for the longer duration of pain in disadvantaged children prior to first clinic review in what has been termed ‘two-tier queueing’ within the Irish system, in that patients with insurance and higher income may be seen more quickly in primary and secondary care at earlier points in a referral pathway. This difference in the utilization of healthcare by families from lower socioeconomic groups has previously been described^ix^.

It is also worth noting the similarities that existed across deprivation groups. Pain intensity, age, sex and parental estimates of their child’s pain and ability to cope with pain were not significantly different across deprivation groups. Outcome measures of pain-related disability such as school attendance, interference with social activities and pain-related sleep disruption were also similar across deprivation groups. Potential limitations include reliance on self-reporting and the sleep disruption question only addressed proportion of disruptions related to pain. The sleep questions in our intake form are loosely based on domains of the Pittsburgh Sleep Quality Index^xv^, addressing sleep hygiene (bedtime and use of screens before bed) and using a similar cut-off (> 60 minutes) for prolonged sleep latency. The rate of screen use before bed in the affluent and average groups is comparable to previously reported rates of 60% in adolescents^xvixvii^. The greater use of screens in lower income families and association with prolonged sleep onset latency have previously been reported and may not be specific to children with pain^xviii^. Given the similar bedtimes and rates of school attendance across deprivation groups, a longer time to sleep onset in the disadvantaged group would imply a shorter sleep duration, although this was not directly questioned. Shorter sleep duration is associated with emotional and behavioral problems, suicidal ideation^xix^ and may precede the onset of depression^xx^. Poor sleep may predispose to pain, pain chronicity and worsen pain-related disability^xxi^. The relationship between poor sleep quality and pain may be mediated worsening low mood and anxiety^xxii^ and greater daytime activity is associated with longer sleep duration in adolescents with chronic pain^xxiii^.

Pain catastrophizing, a construct consisting of rumination, magnification and helplessness, by parents has been identified as influencing pain intensity, pain-related distress and disability in children with chronic pain^xii^ and recent work has suggested that this effect is mediated through child catastrophizing^xxiv^. A reduction in adolescent and parent pain catastrophizing has been associated with improvement in functional disability and parental protective behaviors^xxv^. In adults, associations between lower socioeconomic status and higher levels of pain catastrophizing have been reported in patients undergoing knee surgery^xxvi^. Although not previously described, our finding of higher levels of parental catastrophizing in the disadvantaged group may align with our other findings and related observations in other studies. For example, the scores for the magnification subscale were not significantly different across deprivation groups, which would intuitively fit with parental estimates of child pain intensity and pain coping also being similar across groups. The most significant difference was in the helplessness subscale. One possible explanation for this is the diagnostic uncertainty faced by children and their parents^xxvii^, especially prior to first review at a chronic pain clinic.

Children awaiting review frequently may have increased emergency department attendances and emergency admissions in the years leading up to review, which then abate after pain clinic attendance^xxviii^. Despite clinical need, patients from lower socioeconomic groups tend to underutilize secondary carex^xix^, therefore the first pain clinic appointment may be coinciding with what may have been a more prolonged period of stress and uncertainty in environments that are less familiar to families from lower socioeconomic groups. Ideally, repeated measures of catastrophizing following the initial review would aid in assessing the longitudinal influence on treatment response.

Although essentially from a single centre, this is a national sample with a normal distribution of deprivation scores. As the HP Deprivation Index is specific to Ireland, and although relevant for integrating healthcare with education and housing in an Irish context, it may not be generalizable to other countries. A pain-free control group and longitudinal sampling would also help determine the mediating effects in the relationships between sleep, pain catastrophizing and pain, pain-related disability and outcomes. Additional measures of parental protective behaviors, child pain catastrophizing and a more detailed and validated sleep quality questionnaire would aid in model-building to reproduce and generalize these findings.

## Conclusions

Referrals to the national paediatric chronic pain service in Ireland were normally distributed across social deprivation grades. Children from the disadvantaged group had a longer duration of pain by the time of first clinic review and were more likely to use screens before bedtime and have pronged sleep onset latency. Parents from the disadvantaged group had higher levels of pain catastrophizing. These findings indicate the need to screen for these potential mediators of treatment outcome, which may then require targeted interventions in this group.

## Data Availability

The data used to support the findings of this study are available from the corresponding author upon request.

